# Natural Trajectory of Recovery of COVID-19 Associated Olfactory Loss

**DOI:** 10.1101/2022.02.17.22270551

**Authors:** Amish M. Khan, Jake Lee, Thue Rammaha, Shruti Gupta, Harrison Smith, Thomas Kannampallil, Nyssa Farrell, Dorina Kallogjeri, Jay F. Piccirillo

## Abstract

**Importance:** Prevalence of post-viral olfactory loss has increased dramatically due to the frequency and severity of olfactory dysfunction associated with infection by the SARS-CoV-2 virus.

**Objective:** To determine the trajectory of COVID-19 olfactory loss over a six-month period. A key secondary objective is to assess predictive factors associated with the recovery of olfaction.

**Design:** Longitudinal repeated-measures study that enrolled from May 5, 2020 to February 2, 2021, with the last date of data collection on June 15, 2021.

**Setting:** Barnes-Jewish HealthCare/Washington University School of Medicine facilities (Saint Louis, Missouri, USA).

**Participants:** Individuals who tested positive for SARS-CoV-2 by real-time polymerase chain reaction on nasopharyngeal swab and indicated olfactory loss on COVID-19 screening questionnaire. Individuals were excluded if they had previously diagnosed history of olfactory loss, neurodegenerative disorders, less than 18 years of age, admitted to hospital service, unable to read, write, and understand English, or lacked computer or internet access.

**Interventions/Exposures:** Watch and wait for spontaneous recovery.

**Main Outcome(s) and Measure(s):** Participants completed olfactory assessments every 30 days for six months. Each assessment consisted of the University of Pennsylvania Smell Identification Test (UPSIT), an objective “scratch-and-sniff” test, and Clinical Global Impressions (CGI), a subjective Likert rating scale.

**Results:** The mean age was 41 years old (SD = 16). 39 (80%) were female and 42 (86%) white. At baseline assessment of objective olfaction, 18 (36%) participants had anosmia or severe hyposmia. Subjective, complete recovery at six months was 81% (95% CI 74% to 88%). Likelihood of recovery was associated with age less than 50 years (aHR = 8.1 (95% CI 1.1 to 64.1)) and mild olfactory loss at baseline (UPSIT = 30-33 for males and 31-34 for females) (aHR 6.2 (95% CI 1.2 to 33.0)).

**Conclusions and Relevance:** The trajectory of olfactory recovery among adults with COVID-19 olfactory loss illustrated rapid recovery within 2-3 weeks of infection, and by six months 81% had recovered based on self-report. Age less than 50 years old and mild severity of olfactory loss at baseline were associated with increased likelihood of recovery of olfaction. These findings can be used to inform shared decision-making with patients.

## Introduction

Post-viral olfactory loss is well-documented.^1^ Viruses responsible for olfactory loss include rhinovirus, coronavirus, adenovirus, and influenza virus.^2^ Attention to post-viral olfactory loss has increased dramatically due to the ongoing COVID-19 pandemic, given the frequency and severity of olfactory dysfunction associated with infection by the SARS-CoV-2 virus. In addition, SARS-CoV-2 associated olfactory loss is not accompanied by other upper respiratory infection symptoms, including nasal obstruction or rhinorrhea that is typical with other respiratory viruses causing olfactory loss.^3^

Estimates for the prevalence of acute olfactory loss in COVID-19 infection vary from 34% to 86%.^4–6^ An estimated 44% to 64% of these patients experience recovery of olfaction after two weeks of convalescence from COVID-19 infection.^3,6^ For those with persistent olfactory loss beyond two weeks, it is unknown what percentage of patients continue to have olfactory loss six months from the initial infection.

Loss of sense of smell is associated with decreased quality of life, impaired enjoyment of food, the onset of depressive symptoms, inability to detect harmful environmental hazards including smoke, and reduced social well-being.^7,8^ It has also been linked to increased mortality in older adults.^9^ To date, only a few studies have examined recovery up to six months from initial COVID-19 infection.^10–14^ However, none of these studies assessed subjective and objective measures of olfaction at regular intervals. We describe the trajectory of progression of COVID-19 related olfactory loss over a six-month natural period and assess predictive factors associated with the recovery of olfaction.

## Methods

Candidate study participants were identified through the electronic health record (EHR) within the first two weeks of a positive SARS-CoV-2 test. All study participants were tested for SARS-CoV-2 at Barnes-Jewish HealthCare/Washington University School of Medicine facilities (Saint Louis, Missouri, USA). The study was approved by the Washington University Institutional Review Board (IRB#202004146). Enrollment period was from May 5, 2020 to February 2, 2021, with the last date of data collection on June 15, 2021.

Patients who tested positive for SARS-CoV-2 by real-time polymerase chain reaction on a nasopharyngeal swab and indicated olfactory loss on the COVID-19 screening questionnaire were invited to participate in the study by telephone within two weeks of their positive test. Patients who reported complete return of smell when contacted for participation were included in the number at risk at the time of diagnosis to capture baseline prevalence and recovery from COVID-19 olfactory loss accurately but were not included in subsequent analysis. Individuals were excluded if they had previously diagnosed history of olfactory loss, neurodegenerative disorders (i.e., Alzheimer’s or Parkinson’s disease), were less than 18 years of age, admitted to hospital service, unable to read, write, and understand English, or lacked computer or internet access.

Individuals meeting inclusion criteria were sent an electronic consent form; consenting participants were asked to complete a survey including demographic and clinical information. Enrolled participants were requested to complete olfactory assessments consisting of the *Clinical Global Impression* (CGI)^15^ and *University of Pennsylvania Smell Identification Test* (*UPSIT*; Sensonics, New Jersey).^16^

The *CGI* scale is a subjective Likert rating scale that measures both the severity of dysfunction, *CGI-Severity (CGI-S)*, and rate the improvement (or lack thereof), *CGI-Improvement (CGI-I). CGI-S* is rated from 0-5, with 0 being an absent sense of smell, 3 being good, and 5 being excellent. *CGI-I* is rated from 1-7 with 1 much worse sense of smell, 4 no change, and 7 much better. Subjective, complete recovery was defined by *CGI-S* score of 3 (good), 4 (very good), and 5 (excellent). Subjective, partial recovery was defined by *CGI-I* score of 5 (slightly better), 6 (somewhat better), and 7 (much better).

The *UPSIT* is an objective test consisting of four odor-impregnated booklets that contain ten forced-choice questions each. Participants were asked to self-administer the test by scratching out the odor with a pencil to release the odorants, sniffing the scent, and identifying the odor from four multiple-choice options. Scores are classified into the five clinical categories of normosmia (≥34 for males and ≥35 for females), mild hyposmia (30-33 for males and 31-34 for females), moderate hyposmia (26-29 for males and 26-30 for females), severe hyposmia (19-25), and anosmia (≤18). Objective, complete recovery was defined by UPSIT ≥34 for males and ≥35 for females. Objective, partial recovery was defined as a change in score greater than or equal to 4 points. *UPSIT* has high internal reliability across various populations.^17^

Olfactory assessments were administered every 30 days for up to six months or until participants’ UPSIT scores indicated complete recovery. All participants were instructed to watch and wait for spontaneous recovery; no specific treatment interventions were recommended. Participants self-administered the monthly olfactory assessments and submitted their answers using monthly surveys. Study data were collected and managed using Research Electronic Data Capture (REDCap) electronic data capture system hosted at Washington University.^18,19^ REDCap is a secure, web-based application designed to support data capture for research studies.

Descriptive statistics were used to describe baseline characteristics and assessment scores for all participants. Absolute frequency and relative percentage were used to summarize categorical variables. Chi-square test of independence or Fisher exact test was used to explore the distribution of categorical variables between participants with and without recovery of smell. Mean difference or proportion difference and corresponding 95% confidence intervals (CI) are reported as measures of effect size and precision of estimates. Kaplan-Meier product limit time-to-event analysis was used to explore time to olfaction recovery. The probability of regaining olfaction at different time points was calculated. Time-zero, t(0), was defined as the first abnormal SARS-CoV-2 test. The denominator at t(0) includes individuals who indicated olfactory loss as a symptom of COVID-19, but recovered their sense of smell at the time of telephone call inviting them to participate in the study and individuals who self-reported continued olfactory dysfunction at time of telephone call and agreed to be longitudinally followed. Note t(0.5) denotes two weeks from initial SARS-CoV-2 test and represents the time at which most individuals were contacted by the study coordinators. The denominator at t(0.5) includes individuals who self-reported continued olfactory dysfunction at time of telephone call and agreed to be longitudinally follower. The terminal event was the complete recovery of olfaction defined by *UPSIT* (≥34 for males and ≥35 for females). Participants were censored if they continued to experience an olfactory loss greater than six months in duration or were lost-to-follow-up before six months. Cox-Proportional Hazards regression was used to explore the association of demographic and clinical characteristics with time to olfaction recovery. Proportional hazards assumption was checked using log-log survival plots. Conjunctive consolidation^20^ was used to combine demographic and clinical characteristics to develop a clinical severity staging system to predict likelihood of persistent olfactory dysfunction.

All statistical testing was evaluated with a two-sided test with a pre-specified alpha level of 0.05. Statistical analysis was performed using SPSS (version 28).

## Results

Between April 4, 2020 and January 22, 2021, 325 individuals were identified as having a positive SARS-CoV-2 test and olfactory loss on the COVID-19 screening questionnaire. Among them, 92 individuals indicated that although they had an olfactory loss, they completely recovered their sense of smell by the time of first contact with our research team. A total of 49 individuals self-reported continued olfactory loss at the time of first contact and agreed to participate in the study (Figure 1). The mean age was 41 years old (Standard Deviation (SD)=16), and 31 (63%) participants were less than 50 years old. Of the participants, 39 (80%) were female, and 42 (86%) were White. At baseline *UPSIT* olfactory assessment, 18 (36%) study participants had anosmia or severe hyposmia, 15 (31%) moderate hyposmia, 9 (18%) mild hyposmia, and 7 (14%) tested normal (Table 1). Only 1 (2%) participant reported initiation of treatment for anosmia.

**Table 1.** Baseline Demographic Characteristics of the Study Cohort

| **Characteristics** | **N = 49** |
| --- | --- |
| **Age, n (%)** |  |
| Less than 50 | 31 (63) |
| Greater than or equal to 50 | 18 (37) |
| **Sex, n (%)** |  |
| Female | 39 (80) |
| Male | 10 (20) |
| **Race, n (%)** |  |
| Black | 5 (10) |
| Hispanic | 2 (4) |
| White | 42 (86) |
| **Baseline UPSIT, n (%)** |  |
| Anosmia | 7 (14) |
| Severe Hyposmia | 11 (22) |
| Moderate Hyposmia | 15 (31) |
| Mild Hyposmia | 9 (18) |
| Normosmia | 7 (14) |
| **Number of COVID-19 Symptoms, n (%)** |  |
| 0-3 | 13 (28) |
| 4-6 | 26 (55) |
| 6+ | 8 (17) |
| **Allergic Rhinitis, n (%)** |  |
| Yes | 22 (45) |
| No | 27 (55) |
| **Nasal Trauma or Surgery, n (%)** |  |
| Yes | 2 (4) |
| No | 47 (96) |
| **Chronic Rhinosinusitis, n (%)** |  |
| Yes | 8 (16) |
| No | 41 (84) |
| **Asthma, n (%)** |  |
| Yes | 12 (24) |
| No | 37 (76) |
| **Diabetes Mellitus, n (%)** |  |
| Yes | 7 (14) |
| No | 42 (86) |
| **Fibromyalgia, n (%)** |  |
| Yes | 2 (4) |
| No | 47 (96) |
| **Migraines, n (%)** |  |
| Yes | 8 (16) |
| No | 41 (84) |

**Figure 1.**
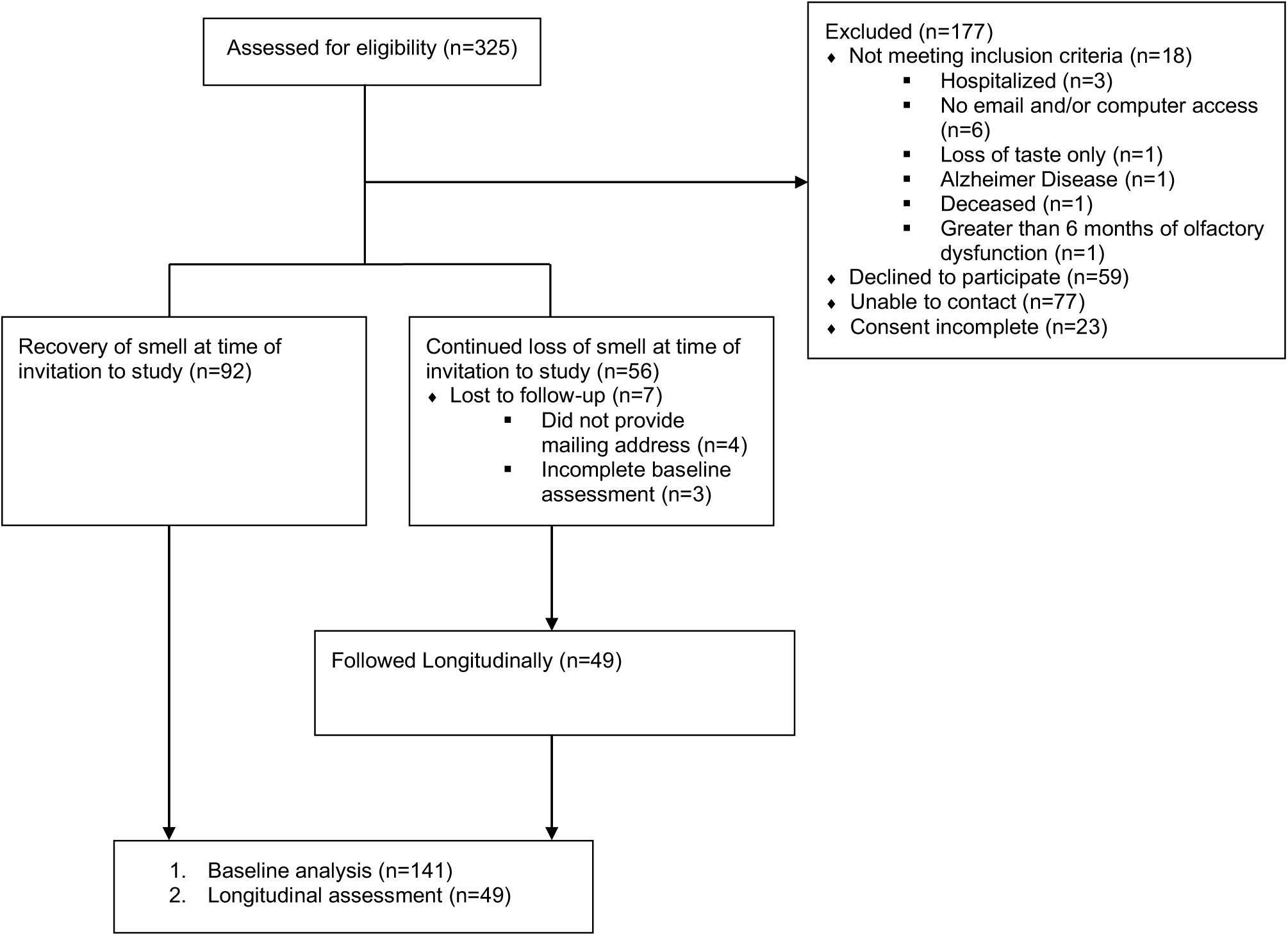
Study Flow Diagram.

There was a rapid recovery in the subjective, self-reported sense of smell in the acute period as defined by *CGI-S* (n = 141). One month from the initial abnormal SARS-CoV-2 test, the recovery rate (i.e., sense of smell described as “good”, “very good”, or “excellent”) was 57% (95% CI 48% to 65%). By the second month, 71% (95% CI 64% to 78%) of study participants reported recovery. Recovery continued at a much slower rate after two months. The probability of self-reported recovery from SARS-CoV-2 resultant olfactory loss by six months was 81% (95% CI 74% to 88%) (Figure 2).

**Figure 2.**
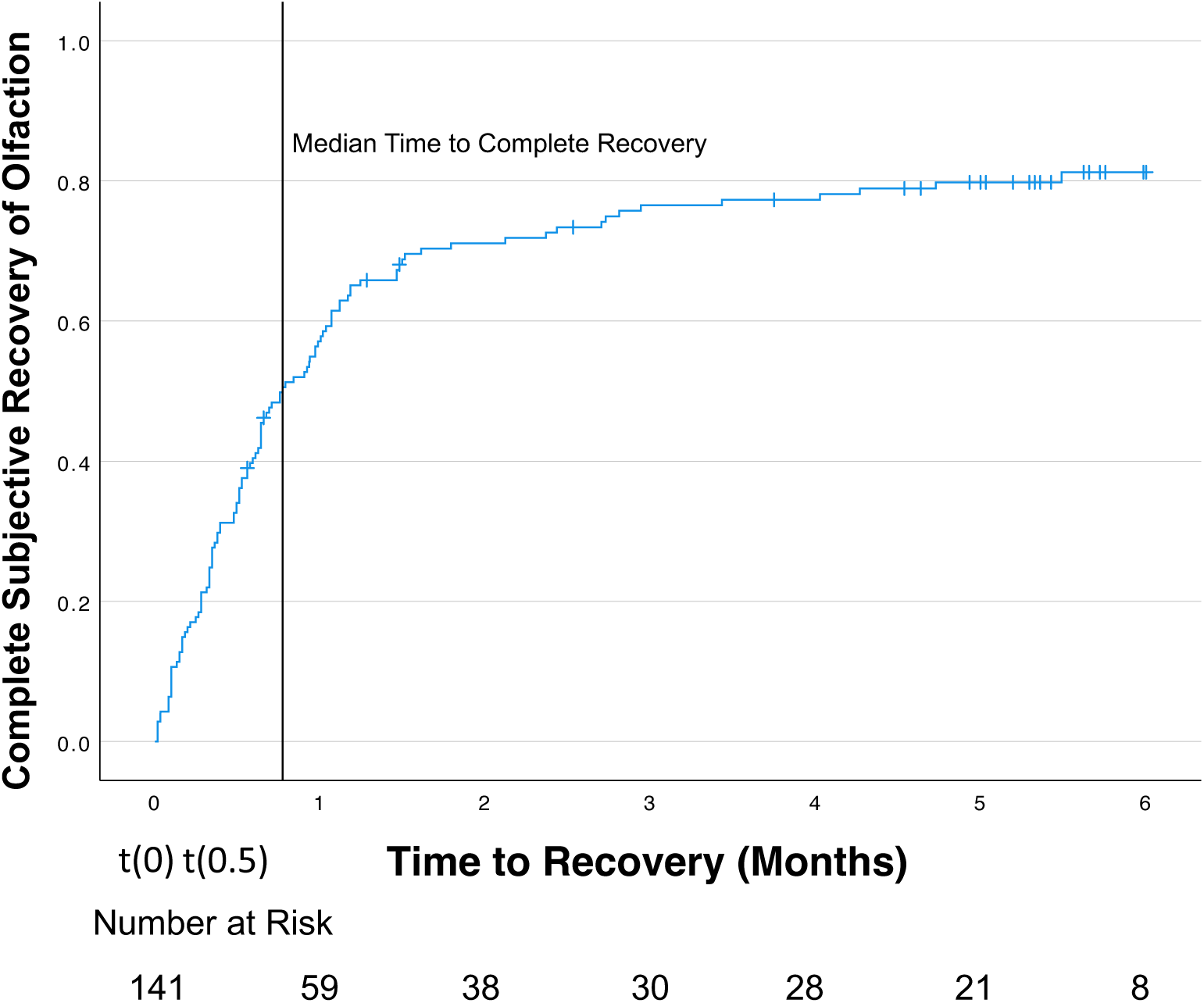
Kaplan Meier Analysis of Complete Subjective Recovery of Olfaction. Kaplan-Meier analysis was used to explore time to recovery of olfaction. Time-zero t(0) is defined as the first abnormal SARS-CoV-2 test. t(0.5) is defined as two weeks from initial SARS-CoV-2 test. Note that 10 individuals who completely recovered sense of smell by the time of the telephone call are included in the denominator beyond t(0.5) because we were unable to contact them within the first two weeks.

Among individuals who continued to have an olfactory loss at the time of study enrollment (n = 49), the probability of subjective, complete recovery as defined by *CGI-S* criterion (good, very good, or excellent [≥ 3]) at two- and six-months was 43% (95% CI 28% to 57%) and 58% (95% CI 43% to 73%), respectively. When partial recovery was defined by *CGI-I* criterion (slightly, somewhat, or much better [≥ 5]), the probability of recovery at two- and six-months was 57% (95% CI 42% to 72%) and 84% (95% CI 72% to 95%), respectively (Figure 3).

**Figure 3.**
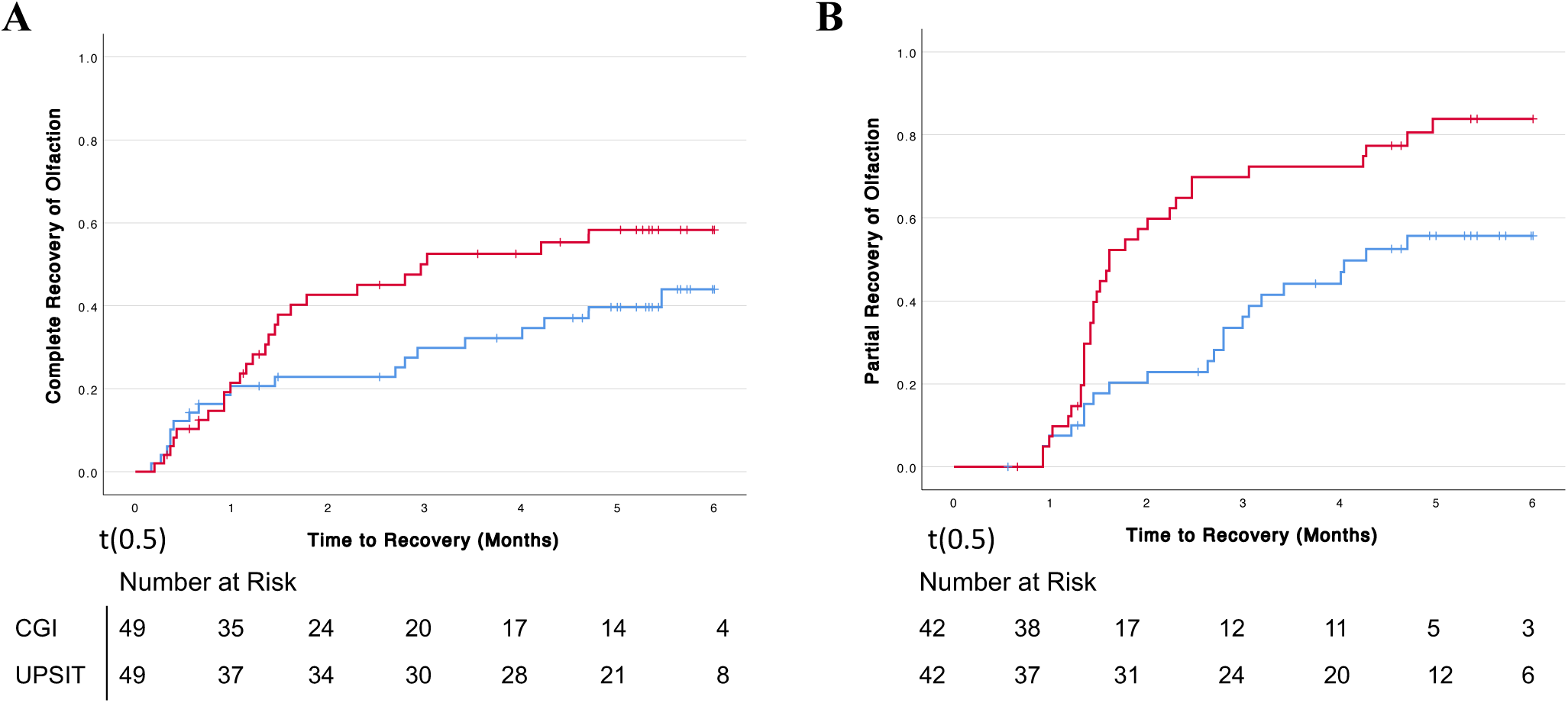
Kaplan-Meier Survival Analysis of Recovery of Olfaction by Various Criteria. Four separate Kaplan-Meier Analyses defining the terminal event by various criterion. t(0.5) is defined as two weeks from initial SARS-CoV-2 test. A: Complete Recovery Complete, objective recovery as defined by *UPSIT* score greater than or equal to 34 and 35 for females and males, respectively, is shown in blue. Complete, subjective recovery as defined by *CGI-S* score of 3 (good), 4 (very good), or 5 (excellent) is shown in red. B: Partial Recovery Partial, objective recovery as defined by *UPSIT* score increase of 4 or greater from baseline assessment is shown in blue. Partial, subjective recovery as defined by *CGI-I* score of 5 (slightly better), 6 (somewhat better), or 7 (much better) is shown in red.

The probability of complete, objective recovery as defined by *UPSIT* ((≥34 for males and ≥35 for females) at two- and six-months follow-up was 23% (95% CI 11% to 35%) and 44% (95% CI 28% to 59%), respectively. The probability of partial recovery defined by *UPSIT* criterion (increase ≥ 4) at two- and six-months was 20% (95% CI 8% to 33%) and 56% (95% CI 40% to 72%), respectively (Figure 3).

As shown in Table 2, there is a gradient in the recovery from olfactory loss across age-group categories and levels of baseline olfactory loss. Next, we used conjunctive consolidation to examine the rates of recovery from olfactory loss within the six conjoined categories of age (<50 years old, >= 50 years old) and baseline olfactory loss (mild hyposmia, moderate hyposmia, severe hyposmia or anosmia) (Table 3). Despite the small numbers in each conjoined cell, the combined prognostic effect of age and baseline severity of smell loss can be demonstrated. For instance, across the two levels of age, there is a gradient for persistent olfactory loss within the levels of baseline olfactory loss. Similarly, across the three levels of baseline olfactory loss, there was a gradient for persistent olfactory loss within each level of age. This association is known as a gradient within a gradient. Cox-Proportional Hazards regression confirmed this relationship between recovery from olfactory loss and age (aHR_Age<50years_ = 8.1 (95% CI 1.1 to 64.1)) and severity of baseline olfactory loss (aHR_Mild Loss_ 6.2 (95% CI 1.2 to 33.0)). To reduce the number of cells and create a clinically useful staging system, conjoint cells with similar persistent olfactory dysfunction rates were consolidated. The new clinical severity staging system for persistent olfactory dysfunction consisted of three categories: Good (2/6, 33%)), Fair (7/13, 54%), and Poor (21/23, 91%). Cox PH regression analysis revealed that relative to the Poor category, the likelihood for recovery of olfactory function for Good is nearly 10 times greater [(HR=10.9 (95% CI 1.9 to 62.6)] and the likelihood for recovery among individuals in the Fair category is nearly 5 times greater [HR=5.5 (95% CI 1.1 to 27.4)].

**Table 2.** Comparison of Baseline Characteristic between Participants with and without Persistent Olfactory Dysfunction by Six Months

| Characteristics | Persistent Olfactory Dysfunction n (%) (n=30) | Absolute Proportions' Difference (95% CI) | Univariate Hazard Ratio for terminal event, recovery of olfaction by UPSIT (95% CI) |
| --- | --- | --- | --- |
| **Age** |  |  |  |
| Less than 50 | 17/31 (55) | 17 | 1.7 (0.6 to 4.8) |
| Greater than or equal to 50 | 13/18 (72) | (-10 to 44) | Ref |
| **Sex** |  |  |  |
| Male | 6/10 (60) | 2 | 1.2 (0.4 to 3.5) |
| Female | 24/39 (62) | (-32 to 36) | Ref |
| **Race** |  |  |  |
| Caucasian | 25/42 (60) | 12 | Ref |
| Non-Caucasian | 5/7 (71) | (-6 to 29) | 1.3 (0.3 to 5.7) |
| **Baseline UPSIT** |  |  |  |
| Mild Hyposmia | 4/9 (44) |  | 5.1 (1.0 to 26.4) |
| Moderate Hyposmia | 10/15 (67) | 22 (8 to 36) | 2.4 (0.5 to 12.4) |
| Anosmia and Severe Hyposmia | 16/18 (89) | 45 (27 to 63) | Ref |
| **Number of COVID-19 Symptoms** |  |  |  |
| 0-5 | 19/34 (56) | 21 | 2.7 (0.8 to 9.3) |
| 6-11 | 10/13 (77) | (-7 to 49) | Ref |
| **COVID-19 Shortness of Breath** |  |  |  |
| Yes | 10/18 (56) | 9 | Ref |
| No | 19/29 (65) | (-20 to 38) | 0.7 (0.3 to 1.7) |
| **COVID-19 Rhinorrhea** |  |  |  |
| No | 20/34 (59) | 10 | 1.6 (0.5 to 4.8) |
| Yes | 9/13 (69) | (-20 to 40) | Ref |
| **Allergic Rhinitis** |  |  |  |
| No | 26/37 (59) | 5 | 0.9 (0.4 to 2.4) |
| Yes | 14/22 (64) | (-20 to 30) | Ref |
| **Chronic Rhinosinusitis** |  |  |  |
| No | 25/41 (61) | 1 | 1.2 (0.3 to 4.0) |
| Yes | 5/8 (62) | (-36 to 38) | Ref |
| **Asthma** |  |  |  |
| No | 21/37 (57) | 18 | 1.5 (0.4 to 5.3) |
| Yes | 9/12 (75) | (-11 to 47) | Ref |
| **Diabetes Mellitus** |  |  |  |
| Yes | 5/7 (71) | 11 | Ref |
| No | 25/42 (60) | (-26 to 48) | 1.3 (0.3 to 5.6) |

**Table 3.** Conjunctive Consolidation of Age and Baseline Olfactory Dysfunction between Participants with and without Persistent of Olfactory Dysfunction at Six Months

| **Baseline Olfactory Dysfunction** | **Age, n (%)** |  |  |
| --- | --- | --- | --- |
|  | < 50 years old | >= 50 years old | Total |
| Mild Hyposmia | 2/6 (33) | 2/3 (67) | 4/9 (44) |
| Moderate Hyposmia | 5/10 (50) | 5/5 (100) | 10/15 (67) |
| Severe Hyposmia or Anosmia | 10/12 (83) | 6/6 (100) | 16/18 (89) |
| Total | 17/28 (61) | 13/14 (93) | 30/42 (71) |

| **Clinical Severity Staging System** |  |
| --- | --- |
| Good | 2/6 (33) |
| Fair | 7/13 (54) |
| Poor | 21/23 (91) |

\*Note: Three individuals < 50 years old and four >= 50 years old tested normosmic at baseline olfactory assessment. We do not know the degree of olfactory dysfunction these individuals had at time-zero because by the time the first UPSIT was delivered to them they had recovered their sense of smell. These individuals are excluded from conjunctive consolidation.

## Discussion

In this study, we found that the majority of individuals recovered quickly from the initial COVID-19 resultant olfactory loss. However, a considerable minority had a persistent loss after three months. This rate of persistent olfactory loss compounded with the high global incidence of COVID-19 infection implies that a large number of infected individuals are likely to develop persistent loss of smell after initial COVID-19 infection. Age and severity of olfactory dysfunction at baseline assessment were significant predictors of the likelihood of persistent loss of smell. These findings can be used to provide patients with appropriate anticipatory counseling and develop patient-centered treatment plans.

The number of individuals with persistent loss identified in this study is comparable to estimates from prior literature, with reported rates of persistent olfactory loss at six months ranging from 4.7% to 27%.^10–12,14,21^ We characterized the trajectory of olfactory function with rapid recovery in the first two months, followed by a slow, incomplete recovery over the following four months. This trend may suggest separate pathophysiology associated with the acute fast recovery and the slow and often incomplete recovery.

Alternatively, this difference in the rate of recovery may be due to differences in baseline risk factors. We found an association between the rate of recovery with age and *UPSIT* score immediately after the initial injury. These data were used to generate a clinical severity staging system by combining age and baseline olfactory loss to discriminate individuals with the highest risk for persistent olfactory loss. The clinical severity staging system can be used to stratify patients at baseline and at the time of randomization for prognostic studies and clinical trials of treatments for COVID-associated anosmia. These findings are consistent with the published literature highlighting an association between persistent olfactory loss and older age.^10,21^ However, these results represent exploratory data analysis, and need to be validated in another dataset. Much more remains to be uncovered about the specific risk factors that predispose COVID-19 patients to continued olfactory loss.

Indeed, the individual trajectories of recovery of our study participants suggest considerable heterogeneity. We identified a mismatch between subjective and objective rates of recovery. Study participants overestimated subjective recovery relative to the level of objective recovery on chemosensory testing. These data suggest that individuals may compensate for their olfactory loss and experience decreased impact on quality-of-life over time. For instance, individuals may cope with their olfactory loss by relying on other senses and invoking cognitive memories of specific scents.^22^ Other individuals may be less bothered by their olfactory loss over time despite continued objective impairment by decoupling their reliance on smell and become less conscious of the deficient overtime.^23^

To our knowledge, only one other study measuring the rate of recovery from COVID-19 olfactory loss at six months has collected intermediate measurements.^10^ In that study, Petrocelli et al. (2021) identified an asymptote at two months beyond which there was little recovery. They suggested that the two-month timepoint is a temporal threshold at which it was reasonable to begin empiric therapy. However, as a caveat, there is no strong evidence supporting the efficacy for most proposed interventions, including oral and intranasal corticosteroids, alpha-lipoic acid, and caroverine.^24,25^ Although we identified a similar inflection point at month two, participants in our cohort had some, albeit slower, rates of recovery beyond this timepoint. This disparity in findings is most likely attributable to baseline differences in the two study cohorts. The authors studied a cohort of patients in whom 47% had anosmia (complete loss of smell) on chemosensory testing at baseline, as compared to 14% in our cohort. We also found that patients with anosmia and severe hyposmia experienced an early plateau in recovery.

This study has the strength of incorporating both validated objective and subjective measures of olfaction. Moreover, participants were identified within two weeks of initial COVID-19 infection, allowing us to accurately capture the initial and six-month recovery of COVID-19 olfactory loss. We collected intermediate measurements at regular intervals each month for six months allowing us to establish the trajectory of improvement over time.

Limitations of this study include the potential for ascertainment bias. Participants who subjectively recovered their sense of smell may have felt less inclined to complete the following month’s olfactory assessment and subsequently be censored in the analysis. Another limitation is the absence of measures of parosmia and phantosmia, which have emerged as major components of COVID-19 olfactory dysfunction. A final limitation is the exclusion of hospitalized patients, who may have a differential rate of recovery from olfactory loss.

It is anticipated that hundreds of thousands of individuals will develop chronic olfactory dysfunction due to COVID-19.^26^ This study provides valuable information about the short-term loss of smell that will allow providers to give accurate anticipatory guidance to patients. We suggest future direction should focus on testing modifications of existing therapies, such as olfactory training, and development of novel therapies for anosmia, such as mindfulness-based stress reduction. Moreover, parosmia and phantosmia have been reported in a high proportion of patients with COVID-19 olfactory dysfunction, but its prevalence has not yet been fully characterized. Lastly, efforts are needed to determine the continued rate of recovery beyond six months.

## Conclusions

In the present study, we defined the trajectory of olfactory recovery among adults presenting with olfactory loss as one of their COVID-19 symptoms. A large majority of patients recovered olfactory function within 2-3 weeks of COVID-19 onset. By six months, 81% had recovered function based on subjective measure with CGI-S. Increasing age and severity of anosmia were associated with reduced likelihood olfaction recovery. Patient-reported assessments of olfactory dysfunction revealed similar patterns of recovery as observed with UPSIT, but generally showed increased rates of recovery at each time point. Results from this study can be used for shared decision-making with patients regarding the likelihood of recovery of olfaction following COVID-19 infection.

## Data Availability

All data produced in the present study are available upon reasonable request to the authors

## Acknowledgements

Tom Naumann at the Washington University School of Medicine Epic1 Research team. Research reported in this publication was supported by the National Center For Advancing Translational Sciences of the National Institutes of Health under Award Number TL1TR002344 (Khan) and the National Institute of Deafness and Other Communication Disorders within the National Institutes of Health, through the “Development of Clinician/Researchers in Academic ENT” training grant, award number T32DC000022 (Gupta, Harrison). The content is solely the responsibility of the authors and does not necessarily represent the official views of the National Institutes of Health (Khan).

---

**Supplementary Table 1.** COVID-19 Symptoms of the Study Cohort

| **Characteristics** | **N = 49** |
| --- | --- |
| **Fever, n (%)** |  |
| Yes | 17 (64) |
| No | 30 (36) |
| **Cough, n (%)** |  |
| Yes | 13 (28) |
| No | 34 (72) |
| **Shortness of Breath, n (%)** |  |
| Yes | 18 (62) |
| No | 29 (38) |
| **Fatigue, n (%)** |  |
| Yes | 21 (45) |
| No | 26 (55) |
| **Muscle or Body Aches, n (%)** |  |
| Yes | 18 (38) |
| No | 29 (62) |
| **Loss of Smell or Taste, n (%)** |  |
| Yes | 49 (100) |
| No | 0 (0) |
| **Sore Throat, n (%)** |  |
| Yes | 19 (40) |
| No | 28 (60) |
| **Rhinorrhea, n (%)** |  |
| Yes | 13 (28) |
| No | 34 (72) |
| **Nausea or Vomiting, n (%)** |  |
| Yes | 7 (15) |
| No | 40 (85) |
| **Diarrhea, n (%)** |  |
| Yes | 11 (23) |
| No | 36 (77) |
| **Headache, n (%)** |  |
| Yes | 16 (34) |
| No | 31 (66) |

